# Demographic benchmarks for equitable coverage of the COVID-19 vaccination program among priority populations

**DOI:** 10.1101/2021.02.18.21251992

**Authors:** Kushagra Vashist, Tabia Akintobi, Robert A. Bednarczyk, KM Venkat Narayan, Shivani A. Patel

## Abstract

We report the demographic distribution of non-institutionalized populations in the US who are prioritized for COVID-19 vaccination by ethnicity, age and sex and region. The composition of non-institutionalized priority populations was estimated using a nationally representative sample of 25,417 adults interviewed in the National Health Interview Survey (NHIS) in 2018. A relatively large fraction of individuals prioritized for the earliest distribution of the vaccine are women, non-Hispanic Black, and young to middle aged adults. Overall, the study provides a platform to track equity in vaccine coverage and to better tailor health communication strategies.

## Background

After nearly 10 months of unprecedented social distancing and over 480,000 American deaths due to COVID-19, 2 SARS-COV-2 vaccines were approved for emergency use in December 2020. Federal and state authorities have developed priorities for vaccination based primarily on protecting individuals who are (1) employed in high-exposure occupations essential to everyday life (2) vulnerable to severe COVID-19 disease (1). There are concerns that differential vaccine coverage and uptake will exacerbate persistent social disparities in COVID-19 infections and outcomes. People of color, especially Black Americans, are at a higher risk of infection, hospitalization and death(2). Yet, reports suggest that public vaccination sites are less likely to be in communities of color in the South (3) and that racial/ethnic minorities are overall more hesitant than whites to take the vaccine(4). This hesitancy may emerge from misinformation, historical or current medical mistreatment, corresponding mistrust and ongoing experiences of discrimination (5). Understanding the demographic composition of the US population prioritized for vaccination is critical to track equity in vaccine coverage and to better tailor health communication strategies.

## Objective

We report the demographic distribution of non-institutionalized populations in the US who are prioritized for COVID-19 vaccination by ethnicity, age and sex and region.

## Methods

Phase 1 and 2 priority populations were defined using the Advisory Committee on Immunization Practices (ACIP) criteria (1). We estimated the composition of non-institutionalized priority populations using a nationally representative sample of 25,417 adults interviewed in the National Health Interview Survey (NHIS) in 2018. These are the most recent national data with appropriate individual-level data on occupation, industry, age, medical conditions, sex, ethnicity, and region of residence. Per the ACIP framework (1), individuals were classified as falling into Phase 1a if they were paid and unpaid healthcare workers who are at a higher risk of direct and indirect exposure and whose work demands physical presence at a healthcare setting. Individuals were classified as being part of Phase 1b if they were non-healthcare essential workers or 75 years and older. Individuals were classified in Phase 1c if they were essential workers not covered in phase 1a or 1b, adults aged 65 to 74, and persons aged 18-64 who had one or more high risk medical conditions for COVID-19. All other individuals were in Phase 2. All analyses were conducted using SAS 9.4 software (SAS Institute Inc., Cary, NC).

## Results

Approximately 85% of US adults fall into Phase 1 and 15% fall into Phase 2 of the vaccine priority schedule (Table). Relative to the US population, Phase 1a has a larger share of women (74%), non-Hispanic Black individuals (18%), and adults aged 25-44 years (41%). Phase 1b has a more equal gender balance (49% women), larger proportion of Non-Hispanic Whites (70%), and by design, adults aged 75 years and older (35%). The demographic distribution of Phase 1c largely follows that of the overall US population. Phase 2 consists of a larger proportion of men (54%), adults aged 18-24 y and 25-44 y, and Hispanic Americans (24%) compared to the overall US population.

**Table.** Distribution of the non-institutionalized adult Covid-19 vaccination priority populations by sex, race/ethnicity, age and region.

|  |  | US Population<br>% (95% CI) | Phase 1<br>% (95% CI) |  |  | Phase 2<br>% (95% CI) |
| --- | --- | --- | --- | --- | --- | --- |
|  |  |  | Phase 1a* | Phase 1b | Phase 1c |  |
| Total number | - | 249,455,533 | 22,074,002 | 54,313,310 | 135,955,691 | 37,112,530 |
| Sex | Men | 48.3<br>(47.5, 49.0) | 26.2<br>(24.0, 28.4) | 51.4<br>(49.9, 52.9) | 49.1<br>(48.1, 50.2) | 53.5<br>(51.3, 55.7) |
|  | Women | 51.7<br>(51.0, 52.5) | 73.8<br>(71.6, 76.0) | 48.6 (47.1,50.1) | 50.9<br>(49.8, 51.9) | 46.5<br>(44.3, 48.7) |
| Race & Ethnicity | Non-Hispanic White | 63.0<br>(61.5, 64.6) | 61.6<br>(58.7, 64.6) | 70.2<br>(68.1, 72.2) | 62.9<br>(61.2, 64.6) | 54.1<br>(51.3, 56.9) |
|  | Non-Hispanic Black | 11.7<br>(10.8, 12.6) | 17.5<br>(15.1, 19.9) | 9.7<br>(8.6, 10.8) | 11.6<br>(10.6, 12.6) | 11.3<br>(9.6, 13.1) |
|  | Hispanic | 16.3<br>(15.0, 17.7) | 10.6<br>(8.7, 12.4) | 13.5<br>(11.8, 15.1) | 16.2<br>(14.8, 17.6) | 24.1<br>(21.6, 26.7) |
|  | Other | 9.0<br>(8.1, 9.8) | 10.3<br>(8.5, 12.1) | 6.6<br>(5.6, 7.6) | 9.3<br>(8.4, 10.2) | 10.5<br>(9.0, 11.8) |
| Age | 18 to 24 | 11.7<br>(11.1, 12.4) | 6.7<br>(5.1, 8.3) | 6.3<br>(5.3, 7.3) | 10.5 (9.7,11.3) | 27.1<br>(25.0, 29.3) |
|  | 25 to 44 | 34.4<br>(33.5, 35.2) | 41.4<br>(38.9, 43.9) | 23.7<br>(22.3, 25.0) | 34.5<br>(33.4, 35.6) | 45.5<br>(43.3, 47.7) |
|  | 45 to 64 | 33.3<br>(32.5, 34.0) | 34.9<br>(32.4, 37.4) | 24.3<br>(23.0, 25.7) | 38.2<br>(37.2, 39.2) | 27.4<br>(25.3, 29.4) |
|  | 65 to 74 | 12.4<br>(11.9, 12.8) | 10.7<br>(9.4, 12.0) | 10.3<br>(9.4, 11.2) | 16.8<br>(16.1, 17.5) | . |
|  | 75 and above | 8.2<br>(7.9, 8.7) | 6.2<br>(5.2, 7.2) | 35.4<br>(33.9, 36.9) | . | . |
| Region | Northeast | 17.3<br>(16.5, 18.2) | 20.9 (18.7, 23.2) | 17.0<br>(15.6, 18.3) | 17.0<br>(16.0, 18.1) | 16.9<br>(15.1, 18.8) |
|  | North Central/Midwest | 22.0<br>(21.1, 22.8) | 21.2<br>(19.2, 23.2) | 24.8<br>(23.3, 26.3) | 21.2<br>(20.2, 22.2) | 21.1<br>(19.1, 23.1) |
|  | South | 36.9<br>(35.8, 38.0) | 37.4<br>(34.8, 40.0) | 35.8<br>(34.0, 37.5) | 37.6<br>(36.2, 39.0) | 35.5<br>(33.0, 37.9) |
|  | West | 23.8<br>(22.7, 24.9) | 20.4<br>(18.4, 22.5) | 22.4<br>(20.8, 24.1) | 24.1<br>(22.9, 25.4) | 26.5<br>(24.0, 28.9) |

\* Comprises essential healthcare workers and does not include institutionalized populations (i.e., long-term care facility residents or incarcerated individuals).
\*\* Comprises individuals 75 years and older and non-healthcare essential workers. Individual criteria for phase 1b does not equal to the total due to overlap between these categories.
\*\*\*Comprises essential workers not covered in phase 1a or 1b, adults aged 65 to 74, and individuals aged 18-64 with medical conditions. Individual criteria for phase 1c does not equal to the total due to overlap between these categories (1).
\*\*\*\*Comprises all individuals not covered in prior phases.
Phases are assigned according to the CDC COVID-19 Vaccination Program Interim Playbook for Jurisdiction Operations framework. See supplementary methods for further detail on the composition of phase 1a, 1b, 1c, and phase 2.
Northeast- New England Division (ME, NH, VT, MA, RI, and CT) and Middle Atlantic Division (NY, NJ, and PA)
North Central/Midwest: East North Central Division (MI, OH, IN, IL, WI) and West North Central Division (MN, IA, MO, ND, SD, KS, and NE)
South: South Atlantic Division (DE, MD, DC, VA, WV, NC, SC, GA, and FL), East South Central Division (KY, TN, MS, and AL), and West South Central Division (TX, AR, OK, and LA)
West: Pacific Division (WA, AK, OR, CA, and HI) and Mountain Division (MT, ID, WY, CO, NM, AZ, UT, and NV)

## Discussion

A relatively large fraction of individuals prioritized for the earliest distribution of the vaccine are women, non-Hispanic Black, and young to middle aged adults. While Black adults are expected to be 17% of Phase 1a, they comprised only 5.4% of adults vaccinated in the first month of the US vaccination program (7). Similarly, the proportion of Black adults vaccinated is lower than their share of the population in 22 of 23 states reporting vaccination data by race (8). Delivering vaccines to priority populations will rely on increased investments in overcoming barriers to vaccine access (such as transportation) and communication strategies that address vaccine hesitancy in vulnerable groups. Beyond national data on vaccination demographics (9), state and county level data to track and address the demographic gap between those who received the COVID-19 vaccine versus those prioritized for vaccination will be critical for an equity-oriented course-correction in the US pandemic response.

## Limitations

Estimates are based on 2018 data. Data constraints imposed the following additional limitations: Phase 1a estimates included all healthcare workers (irrespective of essential classification); estimates exclude individuals in long term care facilities; some Phase 1b and 1c occupations and morbidities were not captured.

## Data Availability

Data used in the manuscrupt: NHIS 2018

https://www.cdc.gov/nchs/nhis/nhis_2018_data_release.htm

## ACKNOWLEDGEMENTS

SAP and KMVN were supported in part by grant number 77624 from the Robert Wood Johnson Foundation. SAP, KMVN, and TA were supported in part by Radx-UP (P30DK111024-05S1). KMVN, TA, and BAB were supported in part by Georgia CEAL (16-312-0217571-66105).

## Supplemental Information

Phase 1a includes critical healthcare workers (Health diagnosing and treating practitioners, health technologists and technicians, other healthcare practitioners and technical occupations, nursing, psychiatric, and home health aides, occupational and physical therapist assistants and aides, other healthcare support occupations, funeral service workers) as per ACIP.

Phase1b includes non-healthcare critical workers (First-line managers/supervisors, protective service workers, firefighting and prevention workers, law enforcement workers, other protective service workers, animal care and service workers, primary, secondary, and special education school teachers, post-secondary teachers, other teachers and instructors, supervisors, farming, fishing, and forestry workers, agricultural workers, fishing and hunting workers, food processing workers, assemblers and fabricators, metal workers and plastic workers, other production occupations, supervisors, production workers, motor vehicle operators, electrical and electronic equipment mechanics, installers and repairers, and woodworkers), adults aged 75 and older not already covered in phase 1a.

Phase1c includes essential workers not previously covered in phase 1a or 1b (Chief executives; general and operations managers; legislators, advertising, marketing, promotions, public relations, and sales managers, operations specialties managers, other management occupations, business operations specialists, financial specialists, mathematical science occupations, architects, surveyors, and cartographers, engineers, drafters, engineering, and mapping technicians, life scientists, physical scientists, social scientists and related workers, Life, physical, and social science technicians, religious workers, counselors, social workers, and other community and social service specialists, lawyers, judges, and related workers, legal support workers, librarians, curators, and archivists, other education, training, and library occupations, media and communication workers, media and communication equipment workers, supervisors, food preparation, and serving workers, cooks and food preparation workers, food and beverage serving workers, other food preparation and serving related workers, supervisors, building and grounds cleaning and maintenance workers, building cleaning and pest control workers, grounds maintenance workers, supervisors, personal care and service workers, transportation and logistics, tourism, and lodging attendants, supervisors, sales workers, retail sales workers, sale representative, services, sale representative, wholesale and manufacturing, other sales and related workers, supervisors, office and administrative workers, communications and IT equipment operators, financial clerks, information and record clerks, material recording, scheduling, dispatching, and distributing workers, secretaries and administrative assistants, other office and administrative support workers, forest, conservation, and logging workers, supervisors, construction and extraction workers, construction trades workers, helpers, construction trades, other construction and related workers, extraction workers, supervisors of installation, maintenance, and repair workers, vehicle and mobile equipment mechanics, installers, and repairers, other installation, maintenance, and repair occupations, printing workers, textile, apparel, and furnishing workers, plant and system operators, supervisors, transportation and material moving workers, air Transportation and logistics workers, rail transportation and logistics workers, water transportation and logistics workers, other transportation and logistics workers, material moving workers), adults aged 65-74 years, and individuals aged 18-65 with one or more of the following comorbidities: Cancer, type 2 diabetes, CKD, COPD, Obesity (BMI>=30), heart condition, asthma, stroke, hypertension, dementia, liver disease as per ACIP.

Phase 2 consists of all workers not covered in prior phases. This includes computer specialists, arts and design workers, entertainers and performers, sports and related workers, entertainment attendants and related workers, personal appearance workers, military and all individuals whose occupations could not be ascertained.

## References

1. Dooling K. The Advisory Committee on Immunization Practices’ Updated Interim Recommendation for Allocation of COVID-19 Vaccine — United States, December 2020. MMWR Morb Mortal Wkly Rep [Internet]. 2021 [cited 2021 Jan 16];69. Available from: https://www.cdc.gov/mmwr/volumes/69/wr/mm695152e2.htm

2. Hawkins D. Differential occupational risk for COVID-19 and other infection exposure according to race and ethnicity. Am J Ind Med. 2020 Sep;63(9):817–20.

3. Across The South COVID-19 Vaccine Sites Missing From Black And Hispanic Neighborhoods [Internet]. NPR.org. [cited 2021 Feb 12]. Available from: https://www.npr.org/2021/02/05/962946721/across-the-south-covid-19-vaccine-sites-missing-from-black-and-hispanic-neighbor

4. Muñana C, 2020. KFF COVID-19 Vaccine Monitor: December 2020 [Internet]. KFF. 2020 [cited 2021 Jan 12]. Available from: https://www.kff.org/coronavirus-covid-19/report/kff-covid-19-vaccine-monitor-december-2020/

5. Khubchandani J, Sharma S, Price JH, Wiblishauser MJ, Sharma M, Webb FJ. COVID-19 Vaccination Hesitancy in the United States: A Rapid National Assessment. J Community Health. 2021 Jan 3;1–8.

6. NewsCAP: COVID-19 vaccine reluctance among U.S. nurses. AJN The American Journal of Nursing. 2020 Nov;120(11):17.

7. Painter EM. Demographic Characteristics of Persons Vaccinated During the First Month of the COVID-19 Vaccination Program — United States, December 14, 2020–January 14, 2021. MMWR Morb Mortal Wkly Rep [Internet]. 2021 [cited 2021 Feb 9];70. Available from: https://www.cdc.gov/mmwr/volumes/70/wr/mm7005e1.htm

8. Pham O, Feb 02 SMP, 2021. Latest Data on COVID-19 Vaccinations Race/Ethnicity [Internet]. KFF. 2021 [cited 2021 Feb 12]. Available from: https://www.kff.org/coronavirus-covid-19/issue-brief/latest-data-covid-19-vaccinations-cases-deaths-race-ethnicity/

9. CDC. COVID Data Tracker [Internet]. Centers for Disease Control and Prevention. 2020 [cited 2021 Feb 12]. Available from: https://covid.cdc.gov/covid-data-tracker

